# Viral shedding and immunological features of children COVID-19 patients

**DOI:** 10.1101/2020.08.25.20181446

**Authors:** Yang Yang, Haixia Zheng, Ling Peng, Jinli Wei, Yanrong Wang, Hexiao Li, Bo Peng, Shisong Fang, Mingxia Zhang, Yanjie Li, Hui Liu, Kai Feng, Li Xing, Jun Wang, Mengli Cao, Fuxiang Wang, Lei Liu, Yingxia Liu, Jing Yuan

**Author notes:** These authors contributed equally to this study (Yang Yang, Haixia Zheng, Ling Peng, Jinli Wei). These authors contributed equally to this study (Yang Yang, Lei Liu, Yingxia Liu, Jing Yuan). **Corresponding author:** Jing Yuan; Yingxia Liu; Lei Liu, and Yang Yang; Shenzhen Third People’s Hospital, 29 Bulan Rd., Shenzhen, 518112, China.

## Abstract

**Background:** SARS-CoV-2 could infect people at all ages, and the viral shedding and immunological features of children COVID-19 patients were analyzed.

**Methods:** Epidemiological information and clinical data were collected from 35 children patients. Viral RNAs in respiratory and fecal samples were detected. Plasma of 11 patients were collected and measured for 48 cytokines.

**Results:** 40% (14/35) of the children COVID-19 patients showed asymptomatic infections, while pneumonia shown by CT scan occurred in most of the cases (32/35, 91.43%). Elevated LDH, AST, CRP, neutropenia, leukopenia, lymphopenia and thrombocytopenia occurred in some cases, and CD4 and CD8 counts were normal. A total of 22 cytokines were significantly higher than the healthy control, and IP-10, IFN-α2 of them in children were significantly lower than the adult patients. Meanwhile, MCP-3, HGF, MIP-1α, and IL-1ra were similar or lower than healthy control, while significantly lower than adult patients. Viral RNAs were detected as early as the first day after illness onset (d.a.o) in both the respiratory and fecal samples. Viral RNAs decreased as the disease progression and mostly became negative in respiratory samples within 18 d.a.o, while maintained relatively stable during the disease progression and still detectable in some cases during 36~42 d.a.o.

**Conclusion:** COVID-19 in children was mild, and asymptomatic infection was common. Immune responses were relatively normal in children COVID-19 patients. Cytokine storm also occurred in children patients, while much weaker than adult patients. Positive rate of viral RNAs in fecal samples was high, and profile of viral shedding were different between respiratory and gastrointestinal tract.

## Background

Coronavirus Disease 2019 (COVID-19) caused by severe acute respiratory syndrome coronavirus 2 (SARS-CoV-2) was firstly reported in late December, 2019 [1, 2], and rapidly caused the ongoing pandemic [3]. As of Apr. 24, 2020, a total of 2,549,632 cases with 175,825 fatal cases were reported all around the world [3]. Although majority of the COVID-19 patients were adults, children patients were also found with a small portion [4-9]. Recently, some studies have revealed the clinical and epidemiological characteristics of children COVID-19 cases, and found that children patients mostly show mild respiratory symptoms and even asymptomatic infection [7, 8, 10].

Viral shedding varies in different viruses and ages, and elucidation of the viral shedding profile is crucial for the laboratory diagnosis, treatment and control of COVID-19. Recent studies have elucidated the viral shedding patterns of SARS-CoV-2 mainly based on adult patients [11-14]. Cytokine storm, which contributes to acute lung injury and development of acute respiratory distress syndrome (ARDS) [15], has been found during SARS-CoV-2 infection [16-18]. Meanwhile, several cytokines have been shown to closely associate with the diseases severity and outcome of COVID-19 in adults [18]. However, whether the cytokine storm occurred in children patients is also currently unclear. In this study, we analyzed the viral shedding and immunological features of children COVID-19 patients based on a case series of 35 laboratory-confirmed children COVID-19 patients.

## Methods

### Patient information and data collection

A total of 35 children COVID-19 patients hospitalized in our hospital were included for the analyses. 50 adult COVID-19 patients and 8 healthy controls reported elsewhere were also included for comparison [18]. Clinical information and laboratory result were collected at the earliest time-point after hospitalization. The study protocol was approved by the Ethics Committees of Shenzhen Third People’s Hospital. Informed consents were obtained from all patients or patients family members. The study was conducted in accordance with the International Conference on Harmonisation Guidelines for Good Clinical Practice and the Declaration of Helsinki and institutional ethics guidelines.

### Quantitative reverse-transcriptase polymerase chain reaction (qRT-PCR)

qRT-PCR were done as previously reported [18, 19]. In brief, throat swabs, nasal swabs, sputum and rectal swabs were collected from children COVID-19 patients upon admission and thereafter. Viral RNAs were extracted from clinical specimens using the QIAamp RNA Viral Kit (Qiagen, Heiden, Germany). A China Food and Drug Administration (CDFA) approved commercially kit (BioGerm Co., Shanghai, China) were used for the detection of SARS-CoV-2 specific RNAs. Samples with a cycle threshold (Ct) value ≤ 37.0 were considered putatively positive. Samples whose Ct was higher than 37 were re-tested and considered positive if Ct was ≥ 37 but ≤ 40 and negative if viral RNAs were undetectable on the second test.

### Cytokine and chemokine measurements

The plasma of the children COVID-19 patients (N=11) were collected at the earliest possible time-point after hospitalization. The concentrations of 48 cytokines and chemokines were measured using Bio-Plex Pro Human Cytokine Screening Panel (Bio-Rad) as previously reported together with plasma collected from 50 adult patients and 8 healthy control [17, 18].

### Disease severity classification of children COVID-19 patients

Disease severity classification were evaluated as previously reported [17, 19], except that the criterion of severe COVID-19 for children is defined as following: 1) respiratory distress with the exclusion of cry and fever (age specific reference ranges: < 2 months: respiration rate (RR) ≥ 60/min; 2~12 months: RR ≥ 50/min; > 5 years: RR ≥ 30/min), 2) resting oxygen saturation ≤ 92%, or 3) Assisted respiration, cyanosis or intermittent apnea, 4) somnolence, convulsions, 5) Refusing to eat or difficult to feed, symptoms of dehydration.

### Statistical analysis

Fisher exact test was used to compare the indicated rates between the symptomatic and asymptomatic cases. Mann-Whitney U test was used to compare plasma cytokine levels between two groups. All statistical tests were calculated using Graphpad Prism 7.0 for Windows. P value less than 0.05 was considered statistically significant.

## Results

### Epidemiological and clinical characteristics of children COVID-19 patients

A total of 35 children COVID-19 patients were included in our study. The median age is 7.5 with a range of 1.5 to 17 years-old, and 13 (37.14%) were male. Two case possessed chronic respiratory diseases, one is asthma and the other pharyngitis. Cough (41.94%) was the most common initial symptoms, followed by fever (38.71%). Expectoration, headache, chill and diarrhea also occasionally happened in some cases. Of note, 40% the cases showed asymptomatic infections. 32 (93.55%) children’s family possessed at least 1 infected family member, while three children were the only COVID-19 cases in their family. 26 children were reported to be either living in Wuhan or having family members who visited Wuhan/Hubei. The median days of onset to admission and duration of hospitalization is 0 and 17, respectively. Viral RNAs were detected in fecal samples from 17 (48.57%) cases. Pneumonia occurred in most of the cases (32/35, 91.43%), and hepatic insufficiency also occurred in some cases (5/35, 14.29%). Of all the 35 cases, 32 cases were moderate, and 3 were mild cases. 34 patients received antiviral treatment within 5 d.a.o, and 85.71% (30/35) of the children received antiviral treatment within 2 d.a.o (Table S1). The combination of interferon alpha and lopinavir was the most frequently used antiviral treatment, and no patients received gamma globulin, corticosteroid or mechanical ventilation treatments. All the patients discharged from hospital (Table S1).

### Laboratory and CT findings of children COVID-19 patients

A complete blood count with differential was assessed for each patient either on the date of hospital admission, or at the earliest time-point after admission (Table 2). Elevated lactate dehydrogenase (LDH), aspartate amino transferase (AST), C-reactive protein (CRP), neutropenia, leukopenia, lymphopenia and thrombocytopenia occurred in some cases with different rates. CT scan of the children patients showed ground-glass, hazy patchy shadows or high density shadows, mainly in the peripheral part of the lung (Figure S1). Moreover, 57.14% of patients showed bilateral involvement of the lung, and 34.29% showed unilateral involvement. Furthermore, based on the initial symptoms, the cases were divided into symptomatic (n=21) and asymptomatic (n=14) groups. Elevated LDH, AST, CRP, neutropenia, leukopenia, lymphopenia as well as unilateral and bilateral involvement of lung were also found in asymptomatic cases, with no statistic differences when compared to the symptomatic cases. Except for CPR which was higher in symptomatic cases, the other indexes of complete blood count were comparable between the two groups.

### Expression profiles of cytokines in children COVID-19 patients

Then, we examined 48 cytokines in the plasma samples, and compared the expression levels with adult COVID-19 patients and healthy adult control. The results showed that 22 cytokines were significantly higher than the healthy control. Of the 22 cytokines, IP-10 and IFN-α2 in children were significantly lower than the adult patients, and the others comparable. Meanwhile, MCP-3, HGF, MIP-1α, IL-6, M-CSF, CTACK and IL-18 were similar with healthy control, while significantly lower than adult patients. IL-1ra was significantly elevated in adult patients, while it was lower than the healthy control in children.

### Profile of viral shedding in children COVID-19 patients

The viral loads were detected by qRT-PCR upon admission and thereafter. Daily measurements of viral load from respiratory and fecal specimens are summarized in Figure 2, lower Ct values indicated higher viral loads. In respiratory samples, high viral loads were found as early as the first day after illness onset (d.a.o), and decreased as the disease progression. Most of patients became negative in respiratory samples within 18 d.a.o, and viral RNAs were re-detectable in two patients at 26, 28 and 33 d.a.o. A total of 17 patients were tested positive in the fecal samples. Viral RNA was also detected as early as the first d.a.o, while viral loads were relatively stable during the disease progression. Viral RNAs were still detectable during 36~42 d.a.o.

**Figure 1.**
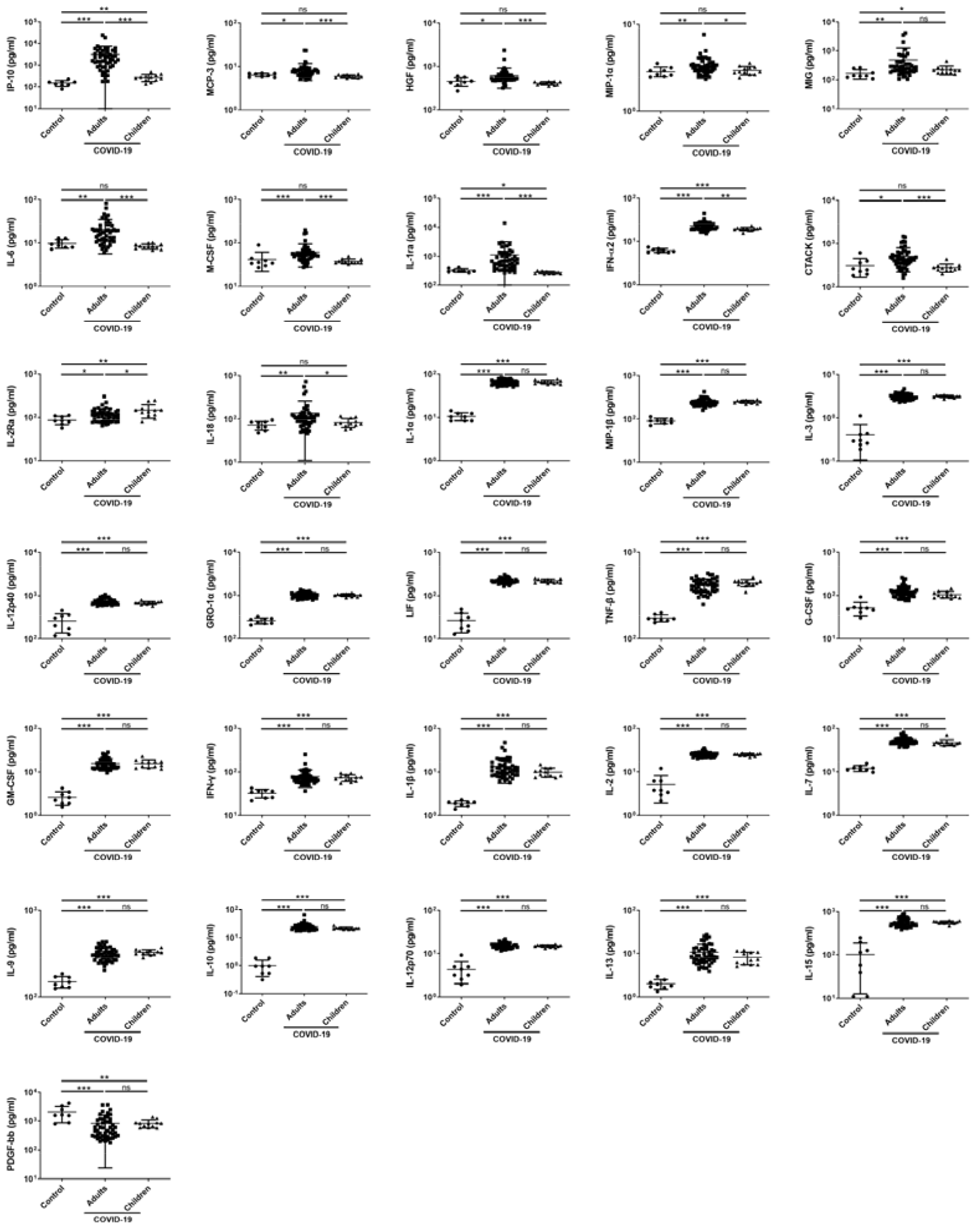
Comparison of serum cytokine concentrations among the children, adult COVID-19 patients and the healthy volunteers. Samples from children (N=11) and adult (N=50) COVID-19 cases were collected at the earliest possible time-point after hospitalization and measured the concentrations of 48 cytokines, and 8 adult healthy subjects were included as control. Values were graphed on a logarithmic scale and presented in units of pg/mL. P values between 0.01-0.05, 0.001-0.01 and 0.0001-0.001 were considered statistically significant (*), very significant (**) and extremely significant (***), respectively, whereas ns represents not significant.

**Figure 2.**
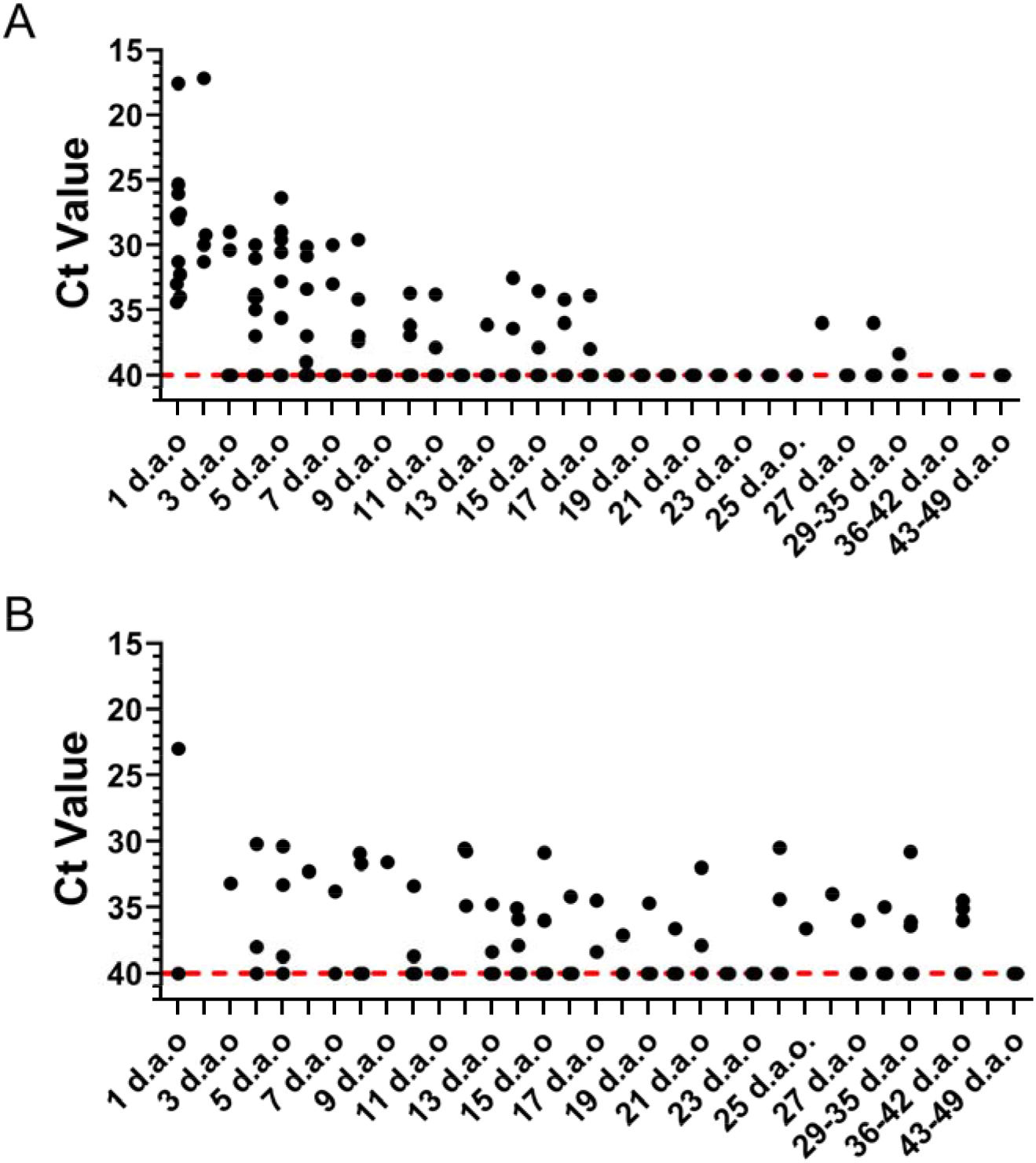
Viral shedding of children COVID-19 patients. Respiratory and fecal samples were collected upon admission and thereafter, and viral RNAs were detected. The obtained Ct values were analyzed in combination with disease progression. The admission date for the asymptomatic patients is defined as the first day after illness onset in this analysis.

## Discussion

Similar with previous study [7], only 37.14% of the children patients showed fever, while cough was most frequent symptoms with a rate of 48.57% (Table 1). A previous study showed that some children COVID-19 cases might display no symptoms or radiologic features of pneumonia, and some might have radiologic features of pneumonia but did not have any symptoms of infection [7]. Notably, 40% of the children cases in our study showed asymptomatic infection before admission (Table 1), however, CT scan of these children showed radiologic features of pneumonia (Table 2). In contrast, 3 of the symptomatic patients showed no radiologic features of pneumonia. All the asymptomatic children cases did the virological tests for investigation, as almost all the patients’ family possessed at least one family member of laboratory confirmed COVID-19 patient. Family cluster of COVID-19 patients were frequently found [10, 20, 21], suggesting that it was easy to transmit among family members, and transmission from family members might serve as the main cause of infection for children. Asymptomatic adult patients were also found with lower rate [4-6], and these patients were also shown to be possible source of infection [22-25]. These asymptomatic patients posed a great threat to the control and prevention of COVID-19, as they were hard to be found unless family members developed symptoms and laboratory confirmed, which is of great concern during the control of COVID-19 pandemic. Meanwhile, there were no statistical differences in epidemiological information, CT scan, blood biochemical and complete blood count between symptomatic and asymptomatic patients (Table 1 and 2), except a higher level of CRP was found in symptomatic patients, which might contribute to the fever of the symptomatic infection.

**Table 1.** Epidemiological and clinical features of children cases hospitalized with SARS-CoV-2 infections.

| Characteristics | SARS-CoV-2 childhood cases |  |  | *P* value |
| --- | --- | --- | --- | --- |
|  | Total (N=35) | Symptomatic (N=21) | Asymptomatic (N=14) |  |
| **Median age (range)** | 7 (1.5, 17) | 8 (1.5, 17) | 7 (1.5, 13) | 0.390 |
| **Age subgroups** |  |  |  |  |
| 0–1 year | 0/35 (0%) | 0/21 (0%) | 0/14 (0%) | 1.000 |
| 1–3 years | 5/35 (14.29%) | 3/21 (14.29%) | 2/14 (14.29%) | 1.000 |
| 3–6 years | 8/35 (22.86%) | 4/21 (19.05%) | 4/14 (28.57%) | 0.511 |
| 6–14 years | 19/35 (54.23%) | 11/21 (52.38%) | 8/14 (57.14%) | 0.782 |
| 14–18 years | 3/35 (8.57%) | 3/21 (14.29%) | 0/14 (0%) | 0.139 |
| **Male (%)** | 13/35 (37.14%) | 9/21 (42.86%) | 4/14 (28.57%) | 0.392 |
| **BMI** | 17.1 (15.0, 19.9) | 17.7 (15.3, 21.1) | 16.3 (13.88, 18.08) | 0.083 |
| **Co-existing chronic medical conditions** |  |  |  |  |
| Chronic heart disease | 0/35 (0%) | 0/21 (0%) | 0/14 (0%) | 1.000 |
| Chronic lung disease | 1/35 (2.86%) | 0/21 (0%) | 1/14 (7.14%) | 0.214 |
| Chronic renal disease | 0/35 (0%) | 0/21 (0%) | 0/14 (0%) | 1.000 |
| Chronic liver disease | 0/35 (0%) | 0/21 (0%) | 0/14 (0%) | 1.000 |
| Diabetes | 0/35 (0%) | 0/21 (0%) | 0/14 (0%) | 1.000 |
| Cancer | 0/35 (0%) | 0/21 (0%) | 0/14 (0%) | 1.000 |
| **Initial symptoms** |  |  |  |  |
| Fever | 13/35 (37.14%) | 13/21 (61.90%) | 0/14 (0%) | <0.0001 |
| Cough | 16/35 (45.71%) | 16/21 (76.19%) | 0/14 (0%) | <0.0001 |
| Expectoration | 1/35 (2.86%) | 1/21 (4.76%) | 0/14 (0%) | 0.407 |
| Headache | 1/35 (2.86%) | 1/21 (4.76%) | 0/14 (0%) | 0.407 |
| Myalgia | 0/35 (0%) | 0/21 (0%) | 0/14 (0%) | 1.000 |
| Chill | 2/35 (5.71%) | 2/21 (9.52%) | 0/14 (0%) | 0.234 |
| Nausea or vomiting | 0/35 (0%) | 0/21 (0%) | 0/14 (0%) | 1.000 |
| Diarrhea | 1/35 (2.86%) | 1/21 (4.76%) | 0/14 (0%) | 0.407 |
| **Disease severity** |  |  |  |  |
| Severe | 0/35 (0%) | 0/21 (0%) | 0/14 (0%) | 1.000 |
| Moderate | 32/35 (91.43%) | 18/21 (85.71%) | 14/14 (100%) | 0.139 |
| Mild | 3/35 (8.57%) | 3/21 (14.29%) | 0/14 (0%) | 0.139 |
| **Exposure history** |  |  |  |  |
| Traveling history to Wuhan/Hubei | 26/35 (74.29%) | 17/21 (80.95%) | 9/14 (64.29%) | 0.269 |
| Family cluster | 32/35 (91.43%) | 19/21 (90.48%) | 13/14 (92.86%) | 0.805 |
| **Anal swab positive of viral RNA** | 17/35 (48.57%) | 5/12 (41.67%) | 8/19 (42.11%) | 1.000 |

| **Interval, median days (IQR)<sup>+</sup>** |  |  |  |  |
| --- | --- | --- | --- | --- |
| Onset to admission | 0 (0, 3) | 0 (0, 3) | NA | NA |
| Admission to discharge | 17 (13, 22) | 17 (13, 22.5) | 16.5 (14, 23.25) | 0.636 |
| **Viral co-infections** | 5/35 (14.29%) | 4/21 (19.05%) | 1/14 (7.14%) | 0.619 |
| **Bacterial co-infections** | 5/35 (14.29%) | 3/21 (14.29%) | 2/14 (14.29%) | 0.531 |
| **Complications** |  |  |  |  |
| Pneumonia | 32/35 (91.43%) | 18/21(85.71%) | 14/14 (100%) | 0.139 |
| ARDS | 0/35 (0%) | 0/21 (0%) | 0/14 (0%) | 1.000 |
| Respiratory failure | 0/35 (0%) | 0/21 (0%) | 0/14 (0%) | 1.000 |
| Hepatic insufficiency | 5/35 (14.29%) | 2/21 (9.52%) | 3/14 (21.43%) | 0.324 |
| Renal insufficiency | 0/35 (0%) | 0/21 (0%) | 0/14 (0%) | 1.000 |
| Cardiac failure | 0/35 (0%) | 0/21 (0%) | 0/14 (0%) | 1.000 |
| Shock | 0/35 (0%) | 0/21 (0%) | 0/14 (0%) | 1.000 |

NA: Not applicable.

**Table 2.** Laboratory and chest radiography findings in children patients of COVID-19 upon admission.

| *Variables* | *Children COVID-19 cases* |  |  | *P values* |
| --- | --- | --- | --- | --- |
|  | **Total (N=35)** | **Symptomatic (N=21)** | **Asymptomatic (N=14)** |  |
| **WBC ($\times 10^9/L$)†** | 5.11 (4.60, 7.79) | 5.11 (4.63, 7.67) | 5.3 (4.37, 7.97) | 0.724 |
| **LYM ($\times 10^9/L$)†** | 2.52 (1.99, 3.27) | 2.49 (1.83, 3.65) | 2.6 (2.18, 2.98) | 0.987 |
| **NEU ($\times 10^9/L$)†** | 2.12 (1.47, 3.53) | 2.25 (1.65, 3.32) | 2.05 (1.35, 3.64) | 0.449 |
| **PLT ($\times 10^9/L$)†** | 253 (222, 298) | 243 (205.5, 309) | 258 (234.75, 299.5) | 0.490 |
| **AST (U/L)†** | 35 (23, 42) | 35 (20.5, 42.8) | 36.45 (22.5, 43.75) | 0.686 |
| **ALT (U/L)†** | 15.9 (11, 24.5) | 16 (11.95, 25.75) | 13.95 (8.93, 24.28) | 0.533 |
| **TB (umol/L)†** | 8.6 (6.1, 11) | 8.8 (5.85, 10.75) | 6.9 (5.65, 11.90) | 0.544 |
| **ALB (g/L)†** | 45.1 (43.5, 46.8) | 45.6 (43.5, 47.25) | 44.4 (43.6, 45.73) | 0.273 |
| **CRE ($\mu\text{mol/L}$)†** | 35 (28, 45) | 35 (26, 48.5) | 31 (28.9, 44.50) | 0.887 |
| **CK (U/L)†** | 83 (68.63, 94.5) | 91.5 (84, 100.25) | 71 (65.25, 92) | 0.113 |
| **LDH (U/L)†** | 261 (198, 496.5) | 266 (224, 516) | 218 (187, 453) | 0.366 |
| **CRP (mg/L)†** | 2.02 (0.52, 5.26) | 4.33 (1.14, 7.85) | 0.75 (0.32, 2.26) | 0.047 |
| **PCT (ng/mL)†** | 0.05 (0.03, 0.08) | 0.06 (0.03, 0.10) | 0.05 (0.01, 0.06) | 0.187 |
| **CD4 (count/$\mu\text{L}$)†** | 781 (684, 931.5) | 857.5 (704.8, 1029.3) | 707 (646, 954) | 0.495 |
| **CD8 (count/$\mu\text{L}$)†** | 600 (512, 922) | 676 (391, 905) | 600 (540, 1062) | 0.558 |
| **CD4/CD8†** | 1.18 (1.06, 1.56) | 1.30 (1.04, 1.58) | 1.09 (1.05, 1.35) | 0.495 |
| **Leukopenia** | 3/35 (8.57%) | 1/21 (4.76%) | 2/14 (14.29%) | 0.324 |
| **Lymphopenia** | 2/35 (5.71%) | 1/21 (4.76%) | 1/14 (7.14%) | 0.766 |
| **Neutropenia** | 8/35 (22.86%) | 3/21 (14.29%) | 5/14 (35.71%) | 0.139 |
| **Neutrophilia** | 0/35 (0%) | 0/21 (0%) | 0/14 (0%) | 1.000 |
| **Thrombocytopenia** | 2/35 (5.71%) | 2/21 (8.33%) | 0/14 (0%) | 0.234 |
| **Elevated AST** | 5/35 (14.29%) | 2/21 (8.33%) | 3/14 (21.43%) | 0.324 |
| **Elevated ALT** | 1/35 (2.86%) | 1/21 (4.76%) | 0/14 (0%) | 0.407 |
| **Elevated CRE** | 0/35 (0%) | 0/21 (0%) | 0/14 (0%) | 1.000 |
| **Elevated CK** | 0/35 (0%) | 0/21 (0%) | 0/14 (0%) | 1.000 |
| **Elevated CRP** | 3/35 (8.57%) | 3/21 (14.29%) | 0/14 (0%) | 0.139 |
| **Elevated LDH** | 12/35 (34.29%) | 8/21 (38.10%) | 4/14 (28.57%) | 0.561 |
| **CT scan** |  |  |  |  |
| Normal | 3/35 (8.57%) | 3/21 (14.29%) | 0/14 (0%) | 0.139 |
| Unilateral involvement | 12/35 (34.29%) | 6/21 (28.57%) | 6/14 (42.86%) | 0.383 |
| Bilateral involvement | 20/35 (57.14%) | 12/21 (57.14%) | 8/14 (57.14%) | 1.000 |

† Values shown represent the mean and inter-quartile range (IQR).

Studies based on the adult COVID-19 patients have showed that viral shedding of SARS-CoV-2 in the respiratory and fecal samples appeared as early as the first two days from illness onset (d.a.o), peaked during 3-5 d.a.o, decreased during diseases progression, and persisted up to 37 d.a.o [12, 13]. Similar with adult patients, viral RNAs could be detected in both the respiratory and fecal samples from children patients, and lasted up to 33 and 41 d.a.o, respectively (Figure 2). Unlike adult patients, viral loads in respiratory samples from children seemed to peaked in the very early stage of disease, usually the first d.a.o, and maintained a middle level during 2-7 d.a.o, then gradually decreased to negative. This observation indicated that viral shedding in children patients might also peak on or before illness onset [13]. Of note, a total of 17 patients (48.57%) were tested positive in the fecal samples, despite that only one patient experienced gastrointestinal symptom. Meanwhile, SARS-CoV-2 shedding in stool is much longer and stable than that in respiratory specimens in children COVID-19 patients (Figure 2). These results highlighted the importance of detecting viral RNAs in fecal samples of children COVID-19 patients. Recently, live virus was successfully isolated from stool [26], and also we isolated the live virus from the rectal swab of a children patient (data not shown), indicating live virus existed in fecal samples. The longer duration of viral shedding in the fecal samples than the respiratory tract (41 vs 33) and the expression of angiotensin-converting enzyme-2 (ACE2) in intestine enterocytes [27, 28], also highlighted the possibility of SARS-CoV-2 infection and replication in the gastrointestinal tract.

Disease severity varied a lot following SARS-CoV-2 infection in adults, from severe ARDS to asymptomatic infections [4-6]. Studies have found that most of the children patients infected with influenza viruses including H5N1, H7N9, and H5N6 avian influenza viruses (AIVs), as well as coronaviruses like SARS-CoV and MERS-CoV showed asymptomatic and mild symptoms [29-34]. In our study, all the included children cases were moderate and mild cases according to the China National Health Commission Guidelines for Diagnosis and Treatment of SARS-CoV-2 infection. Studies have suggested that lymphopenia may associate with disease severity of COVID-19, as 80% of critically ill adult COVID-19 patients experienced lymphopenia, far higher than the adult patients with mild COVID-19 (25%) [5, 6, 35, 36]. Notably, in our study and some other previous report, only a very small portion of children patients experienced lymphopenia [7], which might result from the relatively immature immune system and different immune response compared with adults [37, 38]. This was different from children patients of SARS-CoV infection, which was reported to induce a high portion of lymphopenia up to 46% [34, 37], indicating a different immune response for the two viruses [37]. Another notification is that CD4 and CD8 were normal in children COVID-19 patients (Table 2). However, our previous study has shown that CD4 and CD8 counts were significantly lower in severe and critically ill COVID-19 patients [18], which indicated that the normal CD4 and CD8 counts might also contribute to the mild COVID-19 in children, as CD4 and CD8 cells has been proved to play vital roles in immune responses, including immune regulation, cytokines secretion, virus-specific antibody production and cytolytic activities against target cells [39].

Previous studies have shown that cytokine storm was highly correlated with disease severity and progression [17, 18], and secretion of pro-inflammatory and anti-inflammatory cytokines were found different among different ages of people [38]. So the expression profile of cytokines in children COVID-19 patients upon admission was also analyzed in this study. Similar with adult patients [16-18], elevated concentrations of both pro- and anti-inflammatory cytokines were observed, despite that adult healthy control was used, indicating that cytokine storm also occurred in children following SARS-CoV-2 infection. However, the number of elevated cytokines was far smaller than the adult patients [17, 18]. Furthermore, cytokines which were shown to be highly correlated with disease severity and progression were significantly lower than adult patients, including IP-10, MCP-3, HGF, MIP-1α, and IL-1ra [17, 18]. These results indicated that much weaker cytokine storm happened in children patients, which might also associate with the milder disease in children.

## Conclusions

COVID-19 in children was mild, and asymptomatic infection was common. Immune responses were relatively normal in children COVID-19 patients. Cytokine storm also occurred in children COVID-19 patients, while much weaker than adult patients. Positive rate of viral RNAs in fecal samples was high, and profile of viral shedding were different between respiratory and gastrointestinal tract. To our knowledge, this is the first report on the profile of viral shedding and cytokine expression in children COVID-19 patients, which might provide important information for the understanding of the pathogenesis of SARS-CoV-2 in children.

## Data Availability

The datasets supporting the conclusions of this article are included within the article.

## Abbreviations

COVID-19: Coronavirus Disease 2019
SARS-CoV-2: severe acute respiratory syndrome coronavirus 2
ARDS: acute respiratory distress syndrome
qRT-PCR: Quantitative reverse-transcriptase polymerase chain reaction
CDFA: China Food and Drug Administration
LDH: lactate dehydrogenase
AST: aspartate amino transferase
CRP: C-reactive protein
IL: interleukin
HGF: Hepatocyte growth factor
MCP-3: Monocyte chemotactic protein-3
MIG: Monokine induced gamma interferon
M-CSF: Macrophage colony stimulating factor
G-CSF: Granulocyte colony-stimulating factor
MIP-1α: Macrophage inflammatory protein 1 alpha
CTACK: Cutaneous T-cell-attracting chemokine
IP-10: Interferon gamma induced protein 10

## Declarations

### Ethics approval and consent to participate

The study protocol was approved by the Ethics Committees of Shenzhen Third People’s Hospital. Verbal informed consents were obtained from all patients or patients family members of COVID-19 patients due to the special circumstances that pens and papers were not allowed to be brought into containment facilities.

### Consent to publish

All the authors have read and approved the final version of the manuscript.

### Availability of data and material

The datasets supporting the conclusions of this article are included within the article.

### Authors’ contributions

YY, HZ, LP and JW has equal contribution. JY, ZZ, LL and YY contributed to the study design. HZ, LP, YW, CC, YL, MC, KF, SF, XW and YL contributed to the collection of clinical specimens and data. JW, HZ, MZ, BP, HL and LX performed the quantitative RT-PCR. YY, HZ and JW contributed to the data analysis. YY wrote the manuscript. The corresponding author attests that all listed authors meet authorship criteria and that no others meeting the criteria have been omitted.

## Acknowledgements

We thank the staff of our hospital for their assistance in sample collection and laboratory work.

---

**Figure S1.**
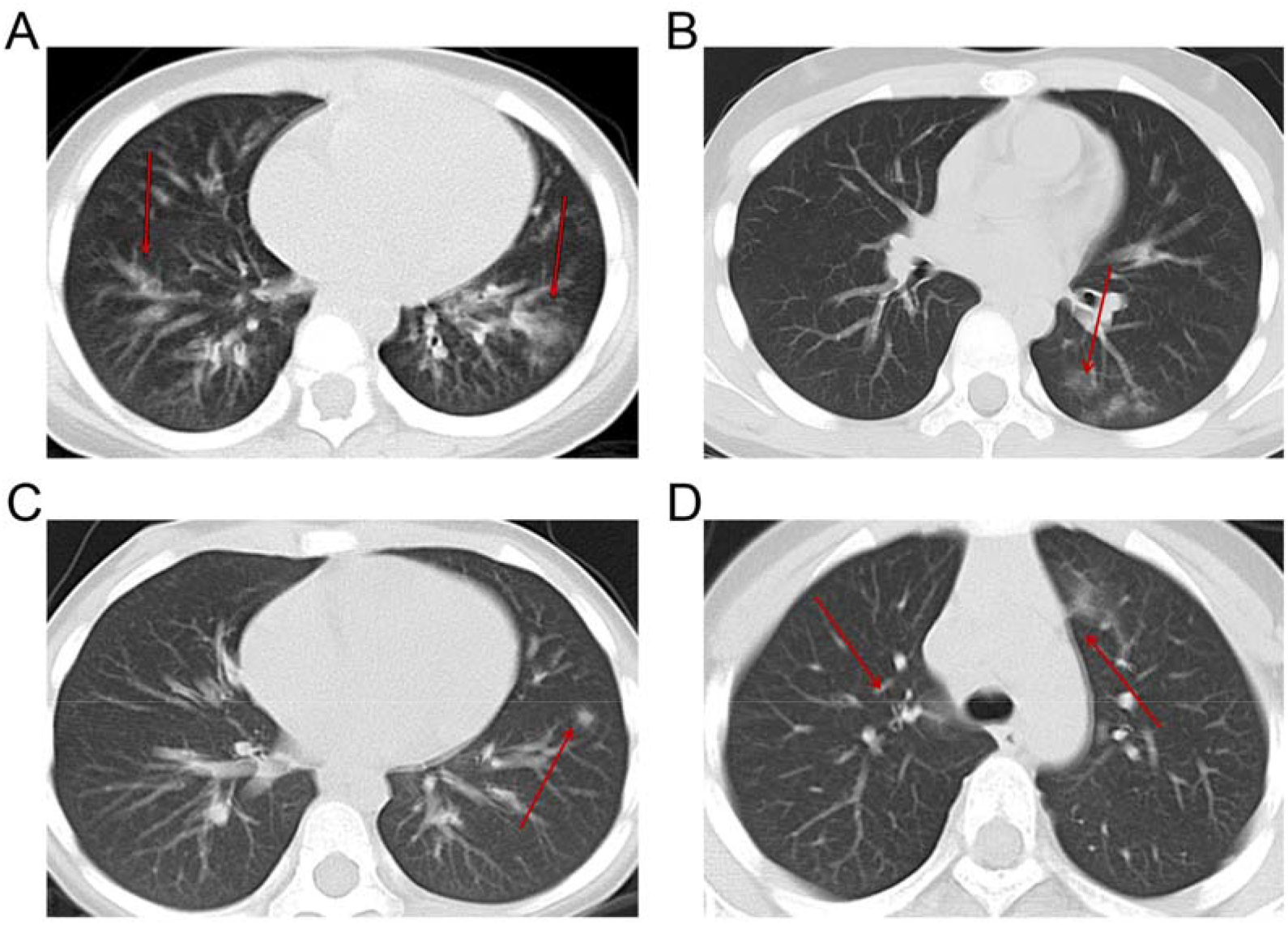
Computed tomography (CT) scan of four children COVID-19 patients. A and B show the CT scan of symptomatic patients with 1 year and 7 months as well as 10 years and 5 months old, respectively. C and D show the CT scan of asymptomatic patients with 3 year and 5 months as well as 7 years and 10 months old, respectively.

**Table S1.** Treatments and outcomes of children COVID-19 patients. Values are numbers (percentages) of patients.

| *Variables* | *Children COVID-19 cases (N=35)* |
| --- | --- |
| **Admission to ICU** | 0/35 (0%) |
| **ARDS** | 0/35 (0%) |
| **Antiviral treatment** |  |
| Interferon alpha | 7/35 (20.00%) |
| Arbidol | 1/35 (2.86%) |
| Interferon alpha + Ribavirin | 2/35 (5.71%) |
| Interferon alpha + Lopinavir | 22/35 (62.86%) |
| Interferon alpha + Lopinavir + Ribavirin | 2/35 (5.71%) |
| Interferon alpha + Lopinavir + Arbidol | 1/35 (2.86%) |
| Received antivirals ≤ 2 days after illness onset | 30/35 (85.71%) |
| Received antivirals 3-5 days after illness onset | 4/35 (11.43%) |
| Received antivirals > 5days after illness onset | 1/35 (2.86%) |
| **Antibiotics** | 0/35 (0%) |
| **Corticosteroid** | 0/35 (0%) |
| **Gamma globulin** | 0/35 (0%) |
| **Outcome** |  |
| Discharge | 35/35 (100%) |
| Death | 0/35(0%) |

ICU: Intensive care unit.
ARDS: Acute respiratory distress syndrome.

## References

1. Zhu N, Zhang D, Wang W, Li X, Yang B, Song J, Zhao X, Huang B, Shi W, Lu R et al. A Novel Coronavirus from Patients with Pneumonia in China, 2019. N Engl J Med 2020.

2. Tan W, Zhao X, Ma X, Wang W, Niu P, Xu W, Gao G, Wu G: A novel coronavirus genome identified in a cluster of pneumonia cases—Wuhan, China 2019– 2020. China CDC Weekly2020, 2(4):61–62.

3. WHO Coronavirus Disease (COVID-19) Dashboard [https://covid19.who.int/]

4. Guan WJ, Ni ZY, Hu Y, Liang WH, Ou CQ, He JX, Liu L, Shan H, Lei CL, Hui DSC et al. Clinical Characteristics of Coronavirus Disease 2019 in China. N Eng J Med 2020.

5. Xu XW, Wu XX, Jiang XG, Xu KJ, Ying LJ, Ma CL, Li SB, Wang HY, Zhang S, Gao HN et al. Clinical findings in a group of patients infected with the 2019 novel coronavirus (SARS-Cov-2) outside of Wuhan, China: retrospective case series. BMJ 2020, 368:m606.

6. Wang D, Hu B, Hu C, Zhu F, Liu X, Zhang J, Wang B, Xiang H, Cheng Z, Xiong Y et al. Clinical Characteristics of 138 Hospitalized Patients With 2019 Novel Coronavirus-Infected Pneumonia in Wuhan, China. JAMA 2020.

7. Lu X, Zhang L, Du H, Zhang J, Li YY, Qu J, Zhang W, Wang Y, Bao S, Li Y et al. SARS-CoV-2 Infection in Children. N Engt J Med2020, 382(17): 1663–1665.

8. Castagnoli R, Votto M, Licari A, Brambilla I, Bruno R, Perlini S, Rovida F, Baldanti F, Marseglia GL: Severe Acute Respiratory Syndrome Coronavirus 2 (SARS-CoV-2) Infection in Children and Adolescents: A Systematic Review. JAMA Pediatr 2020.

9. Tian S, Hu N, Lou J, Chen K, Kang X, Xiang Z, Chen H, Wang D, Liu N, Liu D et al. Characteristics of COVID-19 infection in Beijing. J Infect 2020, 80(4):401–406.

10. Cai J, Xu J, Lin D, Yang Z, Xu L, Qu Z, Zhang Y, Zhang H, Jia R, Liu P et al. A Case Series of children with 2019 novel coronavirus infection: clinical and epidemiological features. Ctin Infect Dis 2020.

11. Zou L, Ruan F, Huang M, Liang L, Huang H, Hong Z, Yu J, Kang M, Song Y, Xia J et al. SARS-CoV-2 Viral Load in Upper Respiratory Specimens of Infected Patients. N Eng J Med 2020, 382(12):1177–1179.

12. Wölfel R, Corman VM, Guggemos W, Seilmaier M, Zange S, Müller MA, Niemeyer D, Jones TC, Vollmar P, Rothe C et al. Virological assessment of hospitalized patients with COVID-2019. Nature 2020.

13. He X, Lau EHY, Wu P, Deng X, Wang J, Hao X, Lau YC, Wong JY, Guan Y, Tan X et al. Temporal dynamics in viral shedding and transmissibility of COVID-19. Nat Med 2020.

14. Yang Y, Yang M, Shen C, Wang F, Yuan J, Li J, Zhang M, Wang Z, Xing L, Wei J: Laboratory diagnosis and monitoring the viral shedding of 2019-nCoV infections. *medRxiv*2020.

15. Channappanavar R, Perlman S: Pathogenic human coronavirus infections: causes and consequences of cytokine storm and immunopathology. Semin Immunopathol 2017, 39(5):529–539.

16. Huang C, Wang Y, Li X, Ren L, Zhao J, Hu Y, Zhang L, Fan G, Xu J, Gu X et al. Clinical features of patients infected with 2019 novel coronavirus in Wuhan, China. Lancet 2020.

17. Liu; Y, Zhang; C, Huang; F, Yang; Y, Wang F, Yuan; J, Zhang Z, Qin; Y, Li; X, Zhao; D et al. 2019-novel coronavirus (2019-nCoV) infections trigger an exaggerated cytokine response aggravating lung injury. Nat! Set Rev 2020.

18. Yang; Y, Shen; C, Li; J, Yuan; J, Wei; J, Huang; F, Li; G, Li; Y, Xing; L, Peng; L et al. Plasma IP-10 and MCP-3 levels are highly associated with disease severity and predict the progression of COVID-19. J Allergy Clin Immunol 2020, 145(4).

19. Liu Y, Yang Y, Zhang C, Huang F, Wang F, Yuan J, Wang Z, Li J, Li J, Feng C et al. Clinical and biochemical indexes from 2019-nCoV infected patients linked to viral loads and lung injury. Set China Life Sei2020, 63(3):364–374.

20. Zhang J, Tian S, Lou J, Chen Y: Familial cluster of COVID-19 infection from an asymptomatic. Crit Care 2020, 24(1):119.

21. Li C, Ji F, Wang L, Wang L, Hao J, Dai M, Liu Y, Pan X, Fu J, Li L et al. Asymptomatic and Human-to-Human Transmission of SARS-CoV-2 in a 2-Family Cluster, Xuzhou, China. Emerg Infect Dis 2020, 26(7).

22. Rothe C, Schunk M, Sothmann P, Bretzel G, Froeschl G, Wallrauch C, Zimmer T, Thiel V, Janke C, Guggemos W et al. Transmission of 2019-nCoV Infection from an Asymptomatic Contact in Germany. N Engl J Med 2020.

23. Jiang X-L, Zhang X-L, Zhao X-N, Li C-B, Lei J, Kou Z-Q, Sun W-K, Hang Y, Gao F, Ji S-X et al. Transmission potential of asymptomatic and paudsymptomatic SARS-CoV-2 infections: a three-family cluster study in China. J infect Dis 2020.

24. Huang L, Zhang X, Zhang X, Wei Z, Zhang L, Xu J, Liang P, Xu Y, Zhang C, Xu A: Rapid asymptomatic transmission of COVID-19 during the incubation period demonstrating strong infectivity in a duster of youngsters aged 16-23 years outside Wuhan and charaderistics of young patients with COVID-19: A prospective contact-tracing study. J infect 2020.

25. Hu Z, Song C, Xu C, Jin G, Chen Y, Xu X, Ma H, Chen W, Lin Y, Zheng Y et al. Clinical characteristics of 24 asymptomatic infections with covid-19 screened among close contacts in Nanjing, China. Sei China Life Sei 2020, 63(1674-7305)706.

26. Yong Z, Cao C, Shuangli Z, Chang S, Dongyan W, Jingdong S, Yang S, Wei Z, Zijian F, Guizhen W et al. Isolation of 2019-nCoV from a Stod Specimen of a Laboratory-Confirmed Case of the Coronavirus Disease 2019 (COVID-19). China CDC Weekly 2020, 2(8): 123–124.

27. Xiao F, Tang M, Zheng X, Liu Y, Li X, Shan H: Evidence for Gastrointestinal Infection of SARS-CoV-2. Gastroenterology 2020.

28. Zhang H, Kang Z, Gong H, Xu D, Wang J, Li Z, Li Z, Cui X, Xiao J, Zhan J et al. Digestive system is a potential route of COVID-19: an analysis of single-cell coexpression pattern of key proteins in viral entry process. Gut 2020.

29. Chakraborty A, Rahman M, Hossain MJ, Khan SU, Haider MS, Sultana R, Ali Rimi N, Islam MS, Haider N, Islam A et al. Mild Respiratory Illness Among Young Children Caused by Highly Pathogenic Avian Influenza A (H5N1) Virus Infection in Dhaka, Bangladesh, 2011. J Infect Dis 2017, 216(suppl_4):S520–S528.

30. Zeng X, Mai W, Shu B, Yi L, Lu J, Song T, Zhong H, Xiao H, Guan D, Wu J et al. Mild influenza A/H7N9 infection among children in Guangdong Province. Pediatr Infect Dis 72015, 34(1):104–107.

31. Chen T, Zhang R: Symptoms seem to be mild in children infected with avian influenza A (H5N6) and other subtypes. Jlnfect20l5, 71(6)702–703.

32. Bi Y, Tan S, Yang Y, Wong G, Zhao M, Zhang Q, Wang Q, Zhao X, Li L, Yuan J et al. Clinical and immunological characteristics of human infections with H5N6 avian influenza virus. Cli Infect Dis 2018.

33. Memish ZA, Al-Tawfiq JA, Assiri A, AlRabiah FA, Al Hajjar S, Albarrak A, Flemban H, Alhakeem RF, Makhdoom HQ, Alsubaie S et al. Middle East respiratory syndrome coronavirus disease in children. Pediatr Infect Dis J 2014, 33(9):904–906.

34. Leung TF, Wong GWK, Hon KLE, Fok TF: Severe acute respiratory syndrome (SARS) in children: epidemiology, presentation and management. Paediatr Respir Rev 2003, 4(4):334–339.

35. Yang X, Yu Y, Xu J, Shu H, Xia Ja, Liu H, Wu Y, Zhang L, Yu Z, Fang M et al. Clinical course and outcomes of critically ill patients with SARS-CoV-2 pneumonia in Wuhan, China: a single-centered, retrospective, observational study. TLancet Respir Med 2020.

36. Li X, Xu S, Yu M, Wang K, Tao Y, Zhou Y, Shi J, Zhou M, Wu B, Yang Z et al. Risk factors for severity and mortality in adult COVID-19 inpatients in Wuhan. J Allergy Clin immunol 2020.

37. Henry BM, Lippi G, Plebani M: Laboratory abnormalities in children with novel coronavirus disease 2019. Clin Chem Lab Med2020.

38. Simon AK, Hollander GA, McMichael A: Evolution of the immune system in humans from infancy to old age. Proc Bio! Sci2015, 282(1821):20143085.

39. Channappanavar R, Zhao J, Perlman S: T cell-mediated immune response to respiratory coronaviruses. Immunol Res 2014, 59(1-3): 118–128.

